# A contemporary look into spontaneous coronary artery dissection: the SwissSCAD registry

**DOI:** 10.1101/2025.03.27.25324803

**Authors:** Sophie Degrauwe, Gregor Fahrni, Christoph Kaiser, Marion Dupré, Stéphane Cook, Thomas Gillhofer, Marco Roffi, Franz Eberli, Daniel Weilenmann, Matthias Bossard, Dik Heg, Hans Rickli

## Abstract

**Background:** Spontaneous coronary artery dissection (SCAD) is an under-recognized cause of acute coronary syndrome. Data regarding contemporary treatment outcomes remain limited, providing the rationale for the establishment of the SwissSCAD registry.

**Objectives:** The primary objective of the SwissSCAD registry, described in this manuscript, is to address contemporary characteristics, management and in-hospital major adverse cardiac events (MACE; defined as a composite of stroke/transient ischemic attack, reinfarction, repeat revascularization and in-hospital death) of patients presenting with SCAD in Switzerland.

**Methods:** We performed an investigator initiated, multicentre, retrospective and prospective observational study including patients with non-atherosclerotic SCAD in 8 centres in Switzerland. Institutional ethics approval and patient consents were obtained. We recorded baseline demographics, precipitating/predisposing conditions, angiographic features, as well as in-hospital treatment and MACE.

**Results:** From August 2020 to March 2024, 264 patients were enrolled. Mean age was 53.4±10.7 years, 85% were women. Cardiovascular risk factors included hypertension (31%), familial history of myocardial ischaemic disease (31%), hypercholesterolemia (25%), active smoking (23%), and diabetes mellitus (3%). Patients presented with STEMI in 35% and with NST-ACS in 59% of cases. Treatment was conservative in 96% of patients. Percutaneous coronary intervention was performed in 4% of patients, coronary artery bypass grafting was limited to 1 patient. In-hospital MACE occurred in 11% of patients with following distribution: stroke/transient ischemic attack (4%), re-infarction (3%), repeat revascularization (2%), in-hospital death (2%). Dual anti-platelet therapy was prescribed in 55% of patients at discharge.

**Conclusions:** Contemporary SCAD patients are treated almost exclusively conservatively. In-hospital MACE rates are sizable, though in-hospital mortality is low.

## INTRODUCTION

Spontaneous coronary artery dissection (SCAD) still represents an underrecognized entity leading to acute coronary syndromes (ACS), which predominantly affects females. The underlying pathophysiology, a non-iatrogenic, non-traumatic separation of the coronary artery wall layers, was first described based on the autopsy of a young woman in 1931^2^. Until twenty years ago, medical literature on SCAD was limited to case reports. Historically, SCAD was considered a rare cause of peripartum-associated myocardial infarction (MI)^1^. Following observational studies and implementation of national registries, publications on the topic have increased exponentially, shaping contemporary understanding of this condition. It estimated that SCAD accounts for 1-4% of ACS in the general population and up to 30% of MI cases in female patients younger than 50 years of age^1^. Patients affected by SCAD are mainly middle-aged women with low prevalence of traditional cardiovascular risk factors^1^, forming a distinct population compared with patients affected by acute coronary syndromes of atherosclerotic aetiology^3,4^. Intracoronary intravascular imaging studies showcased two mechanisms leading to SCAD: formation of intramural hematoma and, less frequently, disruption of the intima^5^. Both mechanisms can lead to antegrade coronary flow reduction, causing acute coronary syndrome in more than 98% of SCAD cases^1^. Patients presenting to the emergency room with SCAD are at risk for under- and missed diagnosis. Young female patients with little burden of cardiovascular risk factors, presenting chest pain, are prone to be discharged from the hospital without coronary angiogram. On the other hand, SCAD might often not be considered in the differential diagnosis of acute coronary syndrome for patients who are not females in the peripartum^6^. Thus, SCAD should figure among the differential diagnosis for the whole spectrum of patients presenting with ACS, since a missed diagnosis might have enduring implications. Clinical features frequently associated with SCAD are hypertension, migraine, fibromuscular dysplasia, peripartum and menopause^6^. Physical exertion and emotional stress are the most common precipitating factors associated with the occurrence of SCAD^6^. Diagnosis is based on coronary angiogram^7^. The Yip-Saw classification describes three angiographic SCAD types^7^. Providing a systemic approach to SCAD angiographic description and diagnosis, the Yip-Saw classification increased physician awareness regarding SCAD^7^. Prospective data derived from large, dedicated SCAD registries will further help to shape understanding of SCAD and pave the way for randomized trials to allow establishment of clinical practice guidelines and standardize clinical care.

In this perspective, the SwissSCAD registry was established, encompassing 8 centres across Switzerland, in order to further assess factors and outcomes associated with SCAD diagnosis.

## METHODS

The SwissSCAD registry is an investigator initiated, multicentre observational study of patients with SCAD, enrolled consecutively across eight hospitals in Switzerland (Centre Hospitalier Universitaire Vaudois, Hôpital Cantonal de Fribourg, Hôpitaux Universitaires de Genève, Stadtspital Zürich Triemli, Universitätsspital Basel, Kantonsspital Baselland, Kantonsspital St. Gallen, and Luzerner Kantonsspital). The central ethic committee was the Geneva Cantonal Commission on Research Ethics (CCER). The study was approved by the local research ethic committee in each participating centre. All patients provided written informed consent for participation. Recruitment started in august 2020. In order to minimize recruitment bias, participation to the SwissSCAD registry was proposed to all consecutive SCAD patients in the participating centres. The study was registered at ClinicalTrials.gov (NCT 04457544). A total of 182 patients were enrolled prospectively (SCAD diagnosis between August 2020 and March 2024) while 82 patients were enrolled retrospectively (SCAD diagnosis between January 2015 and August 2020). Study enrolment was confirmed after angiographic diagnosis validation by three independent and experienced interventional cardiologists. Any discrepancies in opinion were settled by reaching a consensus. Atherosclerotic or iatrogenic coronary dissection excluded patients from participation in the registry. SCAD lesions were classified according to the Yip-Saw Classification^7^. Total distal occlusion, without angiographic signs of distal embolism, was considered type IV SCAD^8^. SCAD recurrence was defined as de novo recurrent SCAD (in contrast to extension of preexisting SCAD). Coronary segmentation was defined according to the SYNTAX classification^9^. Management of SCAD was performed at the discretion of treating physician. The primary objective of the SwissSCAD registry is to collect and analyze following data from consecutive SCAD patients in Switzerland:

- Clinical baseline characteristics
- Predisposing conditions
- Acute management and medical long-term treatment
- Clinical outcomes

The primary outcome measure is in-hospital MACE. The secondary outcome measure is to establish MACE occurrence and SCAD recurrence rates at 30 days, one year and 5 years. In-hospital MACE was defined as the composite of stroke/transient ischemic attack, reinfarction, repeat revascularization and in-hospital death. Follow-up was performed per protocol as clinical visit or telephone visit as well as by mail or email. Data was collected on electronic case report forms using REDCap, hosted in Switzerland. The study was managed by Geneva University Hospitals. Central data monitoring as well as statistical analyses were performed by the Department of Clinical Research, University of Bern. This article is limited to description of the registry’s primary objective.

## STATISTICAL ANALYSES

Baseline demographic and clinical characteristics were depicted as means with standard deviations (p-value from ANOVA) or counts with percentages (p-value from Fisher’s test or chisquare test). Angiographic and discharge data were summarized as counts with percentages (p-value from Fisher’s test or chisquare test). Patient-level angiographic data were depicted as counts with percentages (p-value from Fisher’s test) for multivessel SCAD. For Saw and TIMI classifications, total number of lesions were analyzed (p-values from Poisson models using as offset the Nr of lesions with SCAD). Statistical analyses were performed using Stata 18.0 software. Missing values were not imputed, sample sizes are reported throughout if lower than the total number of patients.

## RESULTS

Between August 2020 and March 2024, 264 patients were enrolled across 8 participating centres in Switzerland. A total of 182 patients were enrolled prospectively (SCAD diagnosis between August 2020 and March 2024) while 82 patients were enrolled retrospectively (SCAD diagnosis between January 2015 and August 2020). Baseline demographic characteristics are summarized in Table 1. Among the 224 female patients (85% of the total cohort), mean age was 53.4±10.6 years (range 22-82 years), while among the 40 male patients the mean age was 53.5±11.0 years (range 30-95 years). In the overall population, prevalence of cardiovascular risk factors was as follows: hypertension 32%, familial history of coronary artery disease 31%, hypercholesterolemia 25%, active smoking 23%, and diabetes mellitus 3%. There was no statistical difference among female versus male patients in regard to prevalence of cardiovascular risk factors (Supplementary appendix 1). Personal history included 1 to 2 previous SCAD events for 12 patients (5%) (Table 2). For those events, clinical presentation was ST segment elevation myocardial infarction (STEMI) in 64% and non-ST segment myocardial infarction (NSTEMI) in 27%. Angiographic characteristics are summarized in Table 2.

**Table 1.**
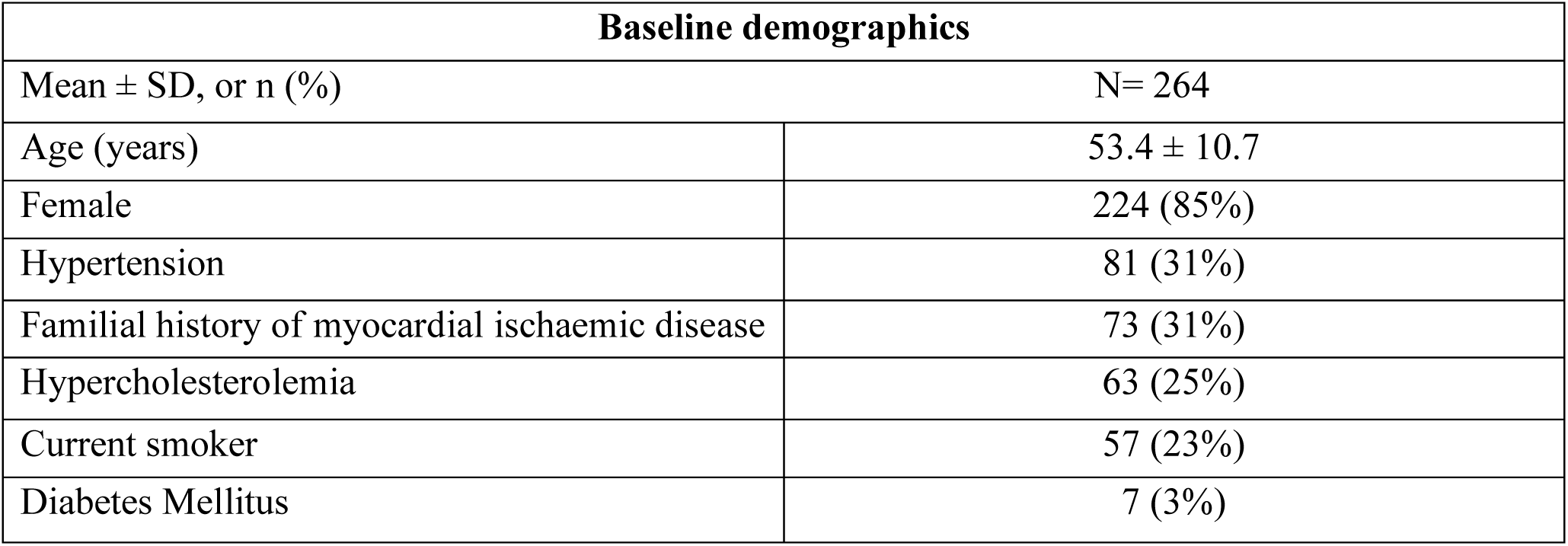
Baseline demographics.

| Baseline demographics |  |
| --- | --- |
| Mean ± SD, or n (%) | N= 264 |
| Age (years) | 53.4 ± 10.7 |
| Female | 224 (85%) |
| Hypertension | 81 (31%) |
| Familial history of myocardial ischaemic disease | 73 (31%) |
| Hypercholesterolemia | 63 (25%) |
| Current smoker | 57 (23%) |
| Diabetes Mellitus | 7 (3%) |

**Table 2.** Previous SCAD events.

| **Previous SCAD events** |  |
| --- | --- |
| Mean ± SD, or n (%) | |
| **Number of previous SCAD events** | N= 12 |
| 1 | 10 (83%) |
| 2 | 2 (17%) |
| **Vessel(s) affected** |  |
| Left anterior descending | 5 (42%) |
| Left circumflex | 5 (42%) |
| Left main | 1 (8%) |
| Intermediate artery | 1 (8%) |
| Marginal of left circumflex | 1 (8%) |
| Right coronary artery | 1 (8%) |
| Posterior interventricular artery | 1 (8%) |
| Retro ventricular artery | 0 (0%) |
| Diagonal artery | 0 (0%) |
| **Clinical presentation** |  |
| STEMI | 7 (64%) |
| NSTEMI | 3 (27%) |
| **Treatment** |  |
| Conservative | 7(64%) |
| Percutaneous coronary intervention | 4 (36%) |
| Coronary artery bypass grafting | 0 (0%) |

SCAD occurrence was associated with one or more triggers in 143 patients (55%) (Table 3), acute emotional stress being the most common one (31%). At admission, 244 patients (92%) reported chest pain, dyspnea was reported in 47 (18%) patients, while 29 patients (11%) complained of nausea or vomiting (Table 3). Most frequent clinical presentations were NSTEMI in 149 patients (57%) and STEMI in 93 patients (35%). The majority of patients had electrocardiogram abnormalities at presentation, namely ST-segment elevation (38%), ST-depression (20%) and T-wave inversion (13%) (Table 3). Peak high-sensitivity cardiac troponin, creatine kinase (CK) and CK-MB were 3745.9±8027.8 ng/l, 793.2 ± 1332.7 U/l, and 98.6 ± 206.9 U/l, respectively (Table 3). Angiographic characteristics are described in Table 4. Single-vessel SCAD was observed in 238 patients (90.2%). The most commonly affected coronary artery was the left anterior descending (LAD) (46%), followed by the left circumflex/marginal branches (LCX) (20%) and the posterior descending artery (10%). Left main coronary artery dissection occurred in 6 patients (2%). On a total of 288 dissections, the most common angiographic SCAD types were type IIa (38.4%) and type IIb (31.5%) followed by type IV (16.6%), type III (6.2%) and type I (7.3%). TIMI flow 3 and TIMI flow 0 were observed in 52.9% and 24.9% of cases respectively. Intracoronary imaging was used in 1 patient. PCI with balloon and stenting was performed in 4 patients (2%). One patient underwent coronary artery bypass grafting. Arterial access for coronary angiography, reported in 103 patients (39% of total cohort), was transradial in 90% of cases (Table 4). Angiographic TIMI flow and angiographic SCAD type per vessel are described in Supplementary Appendix 2.

**Table 3.**
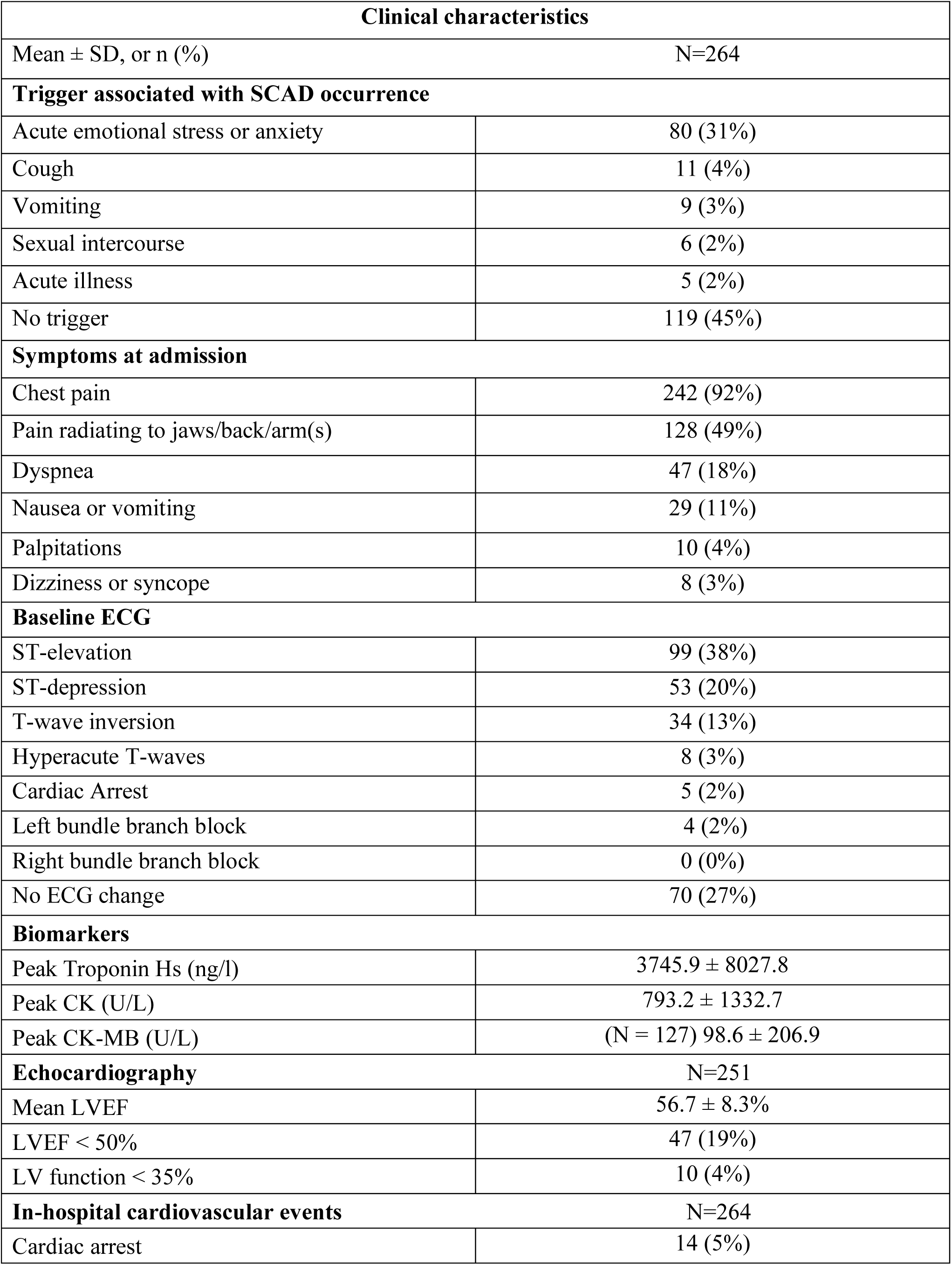
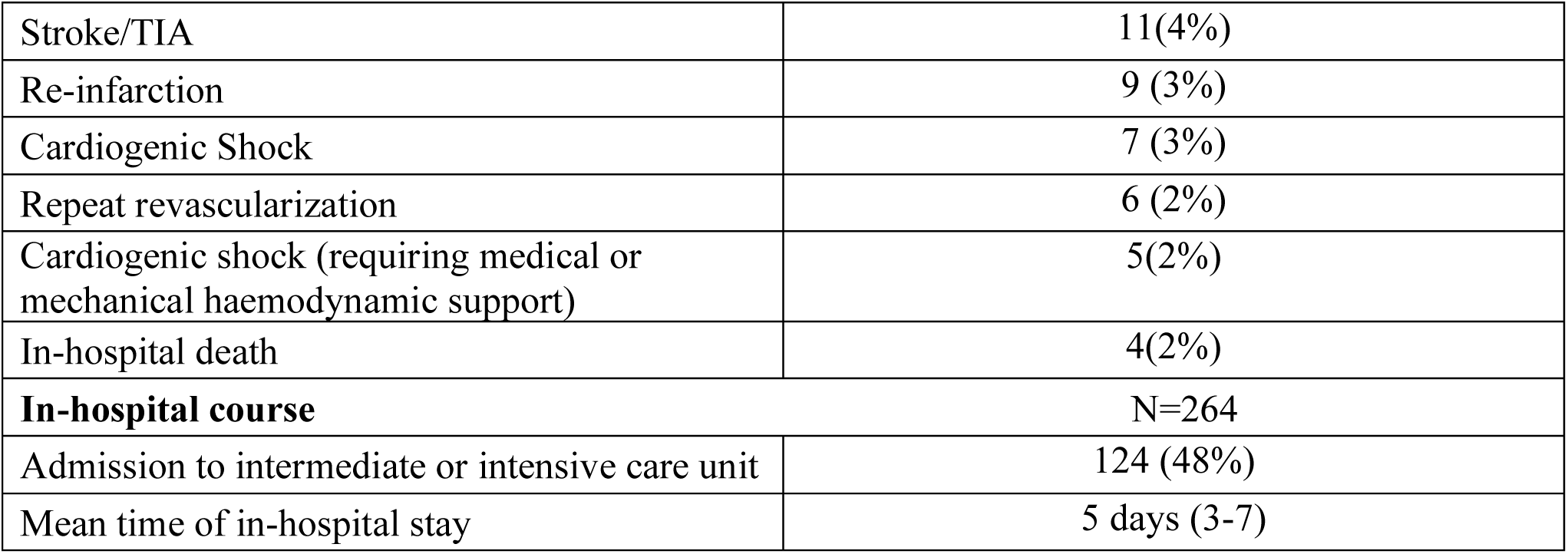
Clinical characteristics.

**Table 4.** Angiographic characteristics.

| **Angiographic characteristics** |  |
| --- | --- |
| Mean ± SD, or n (%) | N=262 |
| Single-vessel SCAD | 237 (90.5%) |
| Multivessel SCAD | 25 (9.5%) |
| **Dissected coronary artery** | N=262 |
| Left anterior descending | 120 (46%) |
| Marginal of left circumflex | 52 (20%) |
| Posterior interventricular artery | 26(10%) |
| Right coronary artery | 23(9%) |
| Left circumflex artery | 21 (8%) |
| Diagonal artery | 20 (8%) |
| Retroventricular artery | 14 (5%) |
| Intermediate artery | 10 (4%) |
| Left main | 6(2%) |
| Septal artery | 3 (1%) |
| **Angiographic SCAD types** | N=288 dissections |
| I | 21 (7.3%) |
| IIa | 111 (38.5%) |
| IIb | 91 (31.6%) |
| III | 18 (6.2%) |
| IV | 47 (16.3%) |
| **Worse TIMI flow** | N=288 dissections |
| 0 | 72 (25.0%) |
| 1 | 34 (11.8%) |
| 2 | 33 (11.5%) |
| 3 | 151 (52.4%) |
| **Arterial access** | N=103 |
| Radial access | 93 (90%) |
| **Invasive treatment** |  |
| PCI with stenting | 4(2%) |
| PCI with plain old balloon angioplasty | 4(2%) |
| CABG | 1(0%) |
| **Repeat coronary angiogram** | N= 55 |
| Number of angiographies per patient |  |
| 1 | 44 (81%) |
| 2 | 8(15%) |
| 3 | 1(2%) |
| 4 | 1(2%) |
| **Indications for repeat coronary angiogram** |  |
| Planned control coronary angiogram | 33 (61%) |
| Suspicion of recurrent SCAD | 14 (26%) |
| Myocardial infarction | 13 (24%) |
| **Invasive treatment** |  |
| PCI with stenting | 7 (13%) |
| PCI with plain old balloon angioplasty | 3 (5%) |
| CABG | 1 (2%) |

Fifty-five patients (20% of total cohort) had more than 1 (2-5) coronary angiogram (Table 4). Indications for repeat coronary angiogram were planned control coronary angiogram (62%), suspicion of recurrent SCAD (25%) and myocardial infarction (24%). During repeat coronary angiogram (Table 4), the most frequently dissected vessels were the LAD (33%), diagonal branches (13%) and RCA (11%). Intracoronary imaging was performed in 2 (4%) patients. PCI with and without stenting were respectively performed in 7(13%) and 3(5%) patients. CABG was performed in 1 patient (Table 4). During index hospitalisation, 126 (48%) patients were admitted to intermediate or intensive care (Table 3). On echocardiogram, mean left ventricular ejection fraction (LVEF) was 57±83%, while 47 patients (19%) and 10 patients (4%) had an LVEF < 50% and < 35%, respectively (Table 3).

With respect to clinical outcomes, in-hospital MACE occurred in 30 patients (11%) with following distribution: stroke/TIA 11 (4%), re-infarction 9 (3%), repeat revascularisation 6 (2%), in-hospital-death 4 (2%) (Table 3). The mean duration of hospital stay was 5 (3-7) days (Table 3).

Regarding registry’s female cohort, comprising 224 individuals, 102 (46%) were post-menopausal (Table 5) and 18 (8%) patients were on hormone replacement therapy for management of menopausal symptoms. At the time of diagnosis, 146 (55%) patients used no contraception. Hormonal or copper coils were the most frequently (16%) used contraceptive methods (Table 5). A minority of female patients (14%) were never pregnant (Table 6). Among patients with pregnancy history, number of pregnancies were distributed as follows: one pregnancy 27 patients (12%), two pregnancies 71 patients (32%), three pregnancies 57 patients (25%), four pregnancies 20 patients (9%), and ≥five pregnancies 12 patients (5%) (Table 5). No patients presented SCAD during pregnancy. A total of 9 (3%) patients presented with SCAD within twelve months following their delivery (Table 5). History of pregnancy complications included pre-eclampsia, eclampsia and HELLP syndrome in 8 (4%) patients, hypertension in 11 (5%) patients, and gestational diabetes in 9 patients (4%) (Table 5).

**Table 5.**
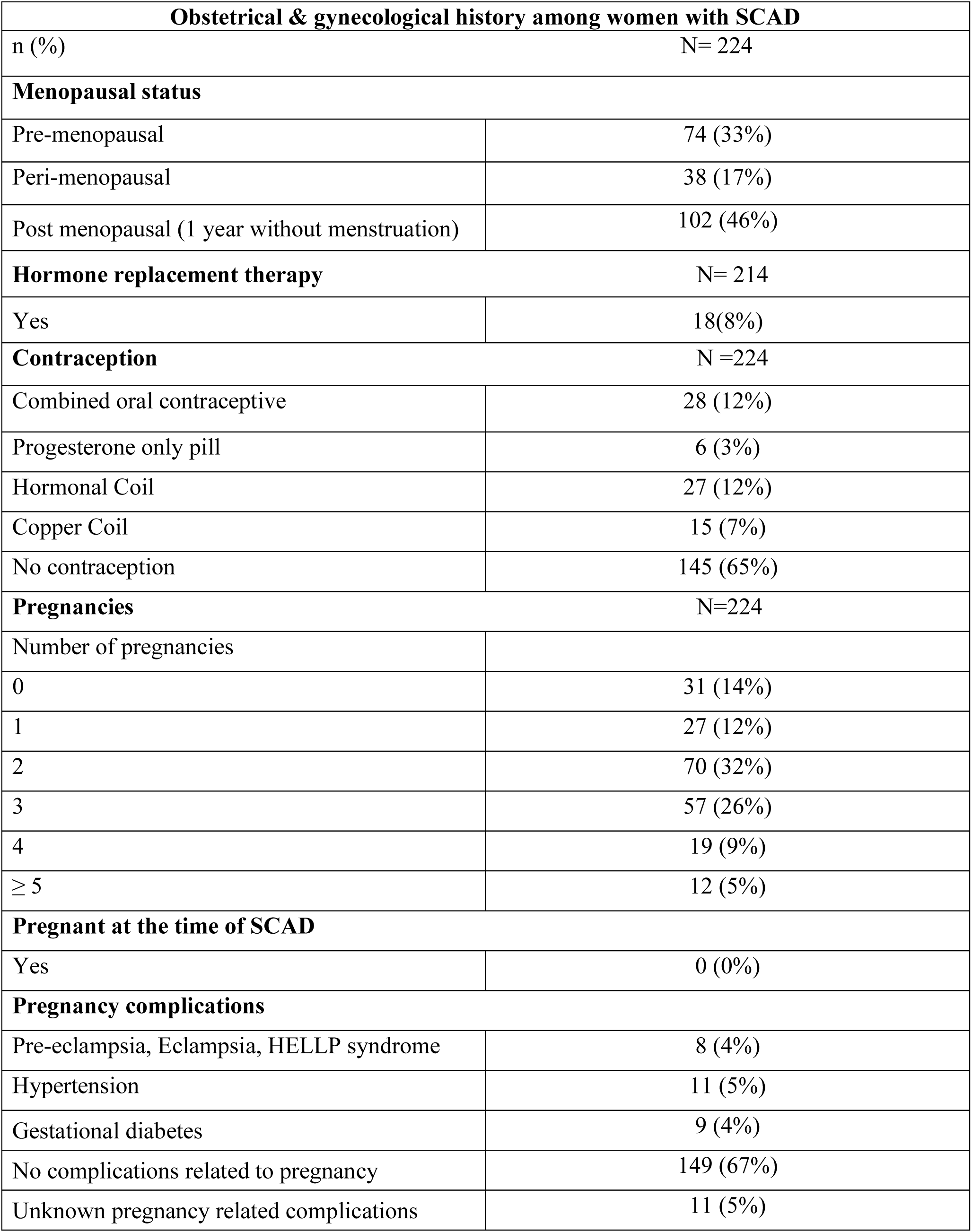
Obstetrical & gynecological history among women with SCAD.

**Table 6.** Medication at discharge.

| **Medication at discharge** |  |
| --- | --- |
| Mean ± SD, or n (%) | N= 264 |
| Aspirin | 229 (87%) |
| DAPT: | 145 (55%) |
| Aspirin and Ticagrelor | 67 (25%) |
| Aspirin and Clopidogrel | 64 (24%) |
| Aspirin and Prasugrel | 14 (5%) |
| Beta blockers | 190 (72%) |
| Angiotensin converting enzyme (ACE) inhibitors or angiotensin II receptor blocker | 160 (61%) |
| Lipid lowering drugs | 13 (51%) |
| Mineralocorticoid receptor antagonists (MRA) | 5 (2%) |

At discharge, aspirin and DAPT were prescribed respectively in 229 (87%) and 145 (55%) patients (Table 6). Beta-blockers, ACE inhibitors or angiotensin II receptor blockers and statins were prescribed at discharge in respectively 72%, 61% and 51% of patients (Table 6).

## DISCUSSION

In this large core laboratory adjudicated multicentre national registry, we present data on baseline characteristics and in-hospital outcomes of SCAD patients enrolled across eight large cardiovascular sites in Switzerland. Participating centres included University Hospitals as well as regional hospitals; situated in the four biggest cities in Switzerland as well as in more rural areas; allowing enrolment of patients coming from varied demographic backgrounds. Core laboratory analysis ensured that all enrolled patients had confirmed angiographic SCAD and were consistently classified.

The cohort age ranges from 22 to 95 years, highlighting the importance of considering SCAD in the differential diagnosis for the entire spectrum of patients presenting with ACS. The cohort was predominantly female (85%) with a mean age of 53.4 years, in line with typical SCAD demographic profiles. Male patients (15%) showed similar age presentation, with no significant differences in cardiovascular risk factors between genders.

The most common cardiovascular risk factors were hypertension (32%), family history of myocardial infarction (31%) and hypercholesterolemia (25%). Diabetes was observed in only 3% of patients, marking a stark contrast with traditional coronary artery disease populations^3^. The prevalence of these risk factors highlights that while SCAD patients often lack classic coronary artery disease risk factors, some overlap exists, particularly in familial predisposition and hypertension. The majority of patients reported a trigger associated to SCAD occurrence, emotional stress being the most frequent one. Recall bias among young and previously healthy individuals may contribute to explain this relatively high frequency of patient-reported precipitants of SCAD, compared to the rates reported in large multinational cohort of patients with ACS^1^. Regardless of this potential overreporting, presenting SCAD adds an additional psychological burden for patients underscoring the need for dedicated patient-centred approaches, such as participation in a dedicated rehabilitation program and peer support.

Consistently with previously published data, patients almost invariably presented with ACS, the majority with NSTEMI while approximately one third had STEMI. High risk clinical presentations such as cardiac arrest, cardiogenic shock, or LVEF < 35% accounted for approximately 10% of the population. Troponin and CK levels showed high variability, attributed to differences in affected coronary artery and segments as well as impact of dissection on coronary antegrade flow. Single-vessel SCAD was noted in 90% of patients. In line with angiographic data from other large SCAD registries, LAD was the most commonly affected coronary artery vessel, suggesting a higher vulnerability for SCAD as compared with other coronary arteries. Intramural hematoma was the predominant form of SCAD in our dataset. Type I SCAD was noted only in 7% of patients. While TIMI flow 3 was observed in a majority of dissections, it is noteworthy that TIMI 0 flow was seen in one fourth of dissected vessels. Our data indicates a very conservative management of SCAD in Switzerland. This seems to distinguish our Swiss cohort from other national SCAD registries^4,15^. Only 3% of patients were treated by coronary revascularization during index hospitalization. In-hospital repeat coronary angiogram was performed in 20% of the patients, mainly driven by “control coronary angiograms”, followed by recurrent myocardial infarction and/or suspicion of recurrent SCAD. On repeat coronary angiogram 10 patients were treated by means of PCI (6 of them were repeat revascularizations), and only 1 patient underwent CABG.

In-hospital survival was excellent (98%), notwithstanding in-hospital MACE occurred in 11% of patients, mainly driven by stroke/TIA, highlighting the importance of in-hospital monitoring after SCAD. In our cohort, 48% of patients were admitted to intermediate or intensive care and mean length of hospitalization was 5 days.

The primary objectives of both short- and long-term medical treatment for SCAD are to relieve symptoms, improve short- and long-term outcomes and reduce the risk of recurrence. There is however a gap of evidence regarding medical therapy following SCAD, current recommendations primarily relying on expert opinions^1,12^. Data from the ongoing BA-SCAD trial will ultimately provide warranted randomized evidence on the safety and efficacy of beta-blocker therapy and dual antiplatelet therapy (DAPT) duration (1 month versus 12 months) in SCAD patients^10^. In the SwissSCAD cohort, patients were almost invariably discharged on aspirin. DAPT was prescribed for 55% patients at discharge. Selection of P2Y12 inhibitor varied across participating centres, ticagrelor and clopidogrel being most frequently prescribed.

According to the European Society of Cardiology and American Heart Association statement papers on SCAD^1,12^, patients undergoing coronary revascularization should receive standard guideline based antiplatelet therapy after PCI^13,14^. Despite the lack of strong evidence supporting the application of DAPT for patients with SCAD treated medically, most experts recommend the usage of aspirin and clopidogrel for 1-12 months^1,12^, followed by long-term aspirin administration^1,12^. Nonetheless, data from a European SCAD registry, call this recommendation into question, since DAPT as compared with SAPT, was independently associated with a higher rate of adverse cardiovascular events at 1-year follow-up^16^.

In light of menorrhagia risk among young premenopausal women, use of antiplatelet therapy among medically managed SCAD patients, should be individualised. Beta-blockers were prescribed in 72% of patients at discharge. LVEF in the cohort suggests that for majority of these patients, beta-blockers were not prescribed according to European Society of Cardiology guidelines for HFrEF of HFmrEF. Rationale for beta-blocker prescription was possibly based on the Vancouver series data demonstrating that use of beta-blockers diminishes (hazard ratio: 0.36; p<0.004) SCAD recurrence^11^. According to European^12^ and American^1^ consensus papers on SCAD, statin therapy should be prescribed according to guideline-based indications for primary and secondary prevention of atherosclerosis, regardless of SCAD event. However, statin prescription is frequent in SCAD cohorts. In the SwissSCAD and Canadian SCAD registries^4^ statins at discharge were prescribed in 51% and 55% of patients, respectively. Reflecting SCADs predilection for middle-aged women, nearly half of female patients were post-menopausal. No patients presented SCAD during pregnancy, precluding reporting of data for this high-risk subset of patients and hinting towards possible underdiagnosis of cardiovascular disease in this group of patients. In line with the French national SCAD registry data^16^, the number of pregnancies per patient was high; 26% of patients reported three pregnancies, 9% of patients reported four pregnancies and 5% of patients reported five or more pregnancies. A small subset of patients presented personal history of pre-eclampsia, eclampsia, gestational diabetes, and pregnancy-related hypertension. Those pregnancy complications are established risk factors for future cardiovascular disease, including cardiovascular mortality; also, after adjusting for confounding factors^17^. Notwithstanding, there is a lack of data on pregnancy complications and their potential link to future SCAD occurrence.

## CONCLUSION

The SwissSCAD registry allows for a contemporary insight into patient characteristics, management and in-hospital outcomes. Key findings underscore SCAD’s predominance in middle-aged women, frequent presentation with NSTEMI and the conservative management chosen for the vast majority of patients. While in-hospital survival was excellent, approximately 1 out of 10 patients suffered an in-hospital MACE, highlighting the importance of in-hospital surveillance after SCAD.

## IMPACT ON DAILY PRACTICE

SCAD predominantly affects middle-aged women and patients almost invariably present with ACS. In-hospital MACE rates are sizable, though in-hospital mortality is low. The SwissSCAD registry provides real-world evidence on in-hospital outcomes of a large SCAD cohort. Contemporary SCAD management is predominantly conservative. In-hospital MACE rates are consistent with those reported in other large national registries. Long-term follow-up of SCAD registries will provide data regarding predictors of adverse clinical outcomes, risk of recurrence and long-term management.

## LIMITATIONS

Our study is non-randomized. All consecutive SCAD patients were approached for enrolment in the registry. However, data regarding patients not surviving before hospital admission and underdiagnosed patients were not captured. No patients presented SCAD during pregnancy, precluding reporting of data for this high-risk subset of patients and hinting towards possible underdiagnosis of cardiovascular disease in this group of patients. Clinical events were not adjudicated by a clinical committee event.

## Data Availability

All data referred to in this manuscript will be made available to the Editor and reviewers if needed.

## ABBREVIATIONS

ACS: Acute coronary syndrome
CABG: Coronary artery bypass graft
DAPT: Dual antiplatelet therapy
LAD: Left anterior descending
LCX: Left circumflex artery
LVEF: Left ventricular ejection fraction
MACE: Major adverse cardiovascular events
MAE: Major adverse events
MI: Myocardial infarction
NSTEMI: Non-ST-elevation myocardial infarction
PCI: Percutaneous coronary intervention
PIA: Posterior interventricular artery
RCA: Right coronary artery
SCAD: Spontaneous coronary artery dissection
STEMI: ST-elevation myocardial infarction
TIA: Transient ischemic attack

---

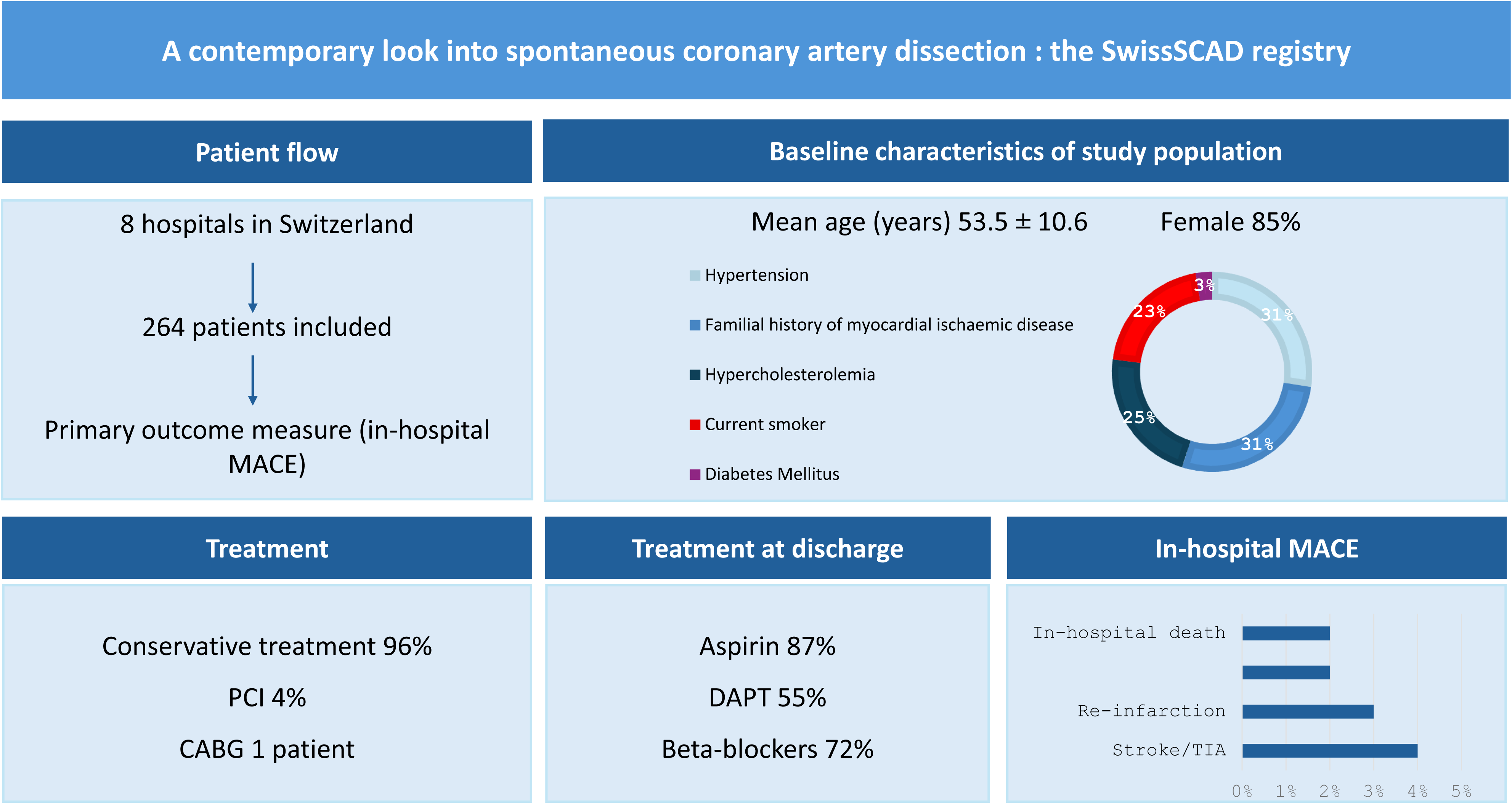

